# Disentangling the roles of trauma and genetics in psychiatric disorders using an Electronic Health Records-based approach

**DOI:** 10.1101/2022.08.28.22279306

**Authors:** Shelby Marchese, Winston Cuddleston, Carina Seah, Jessica Johnson, Laura M. Huckins

## Abstract

Posttraumatic stress disorder (PTSD) requires an exposure to trauma for diagnosis by the DSM-V. Despite this, there is no documented linear relationship between degree of trauma and severity/chronicity of PTSD.

To determine whether traumatic and stressful life events (TSLEs) collected from Electronic Health Records (EHR) interact with PTSD genetics to better define individual risk of developing PTSD. We collected trauma information from patient records in the Mount Sinai Bio*Me*™ biobank population-based cohort and tested for associations with PTSD. We generated a TSLE risk score (TRS), tested its association with PTSD, and for interactions with a polygenic risk score (PRS) of PTSD and a GWAS of PTSD using our biobank population.

We used the Mount Sinai Bio*Me*™ biobank patient population of 31,704 individuals with matched genotype and EHR data, which has been enrolling patients since 2006. Patient enrollment in Bio*Me*™ is unrestricted, resulting in high diversity. Our study includes 1,990 individuals with PTSD and 28,478 individuals without PTSD from the Mount Sinai Bio*Me*™ biobank.

A total of 1,990 individuals with PTSD and 28,478 controls were analyzed (average age 42-78 years, 58.7% women). We identified a total of 22 EHR-derived TSLEs that were associated with PTSD (*β*> 0.029, p<1.61×10^−3^). TSLEs interacted with each other to increase the risk of developing PTSD, with the most significant interaction between being female (as a proxy for experiencing sexism) and being HIV+ (*β*=0.013, p=8.9×10^−11^). PRS of PTSD and lead SNPS from GWAS interacted with TSLEs and our TRS to increase PTSD risk. In addition to TRS interactions, we found significant interactions between PTSD PRS and domestic violence, and sexual assault in European Americans (*β*>207.74, p<1.80×10^−3^). rs113282988 and rs189796944 variants reached genome-wide significance in interactions with TRS (*β*>0.056, p<9.04×10^−9^), and rs189796944 in an interaction with Physical Survival TSLEs (*β*>0.127, p<4×10^−8^).

In conclusion, quantification of trauma type, severity, and magnitude—alone and in concert with genetics—provides better prediction of PTSD risk than PRS alone. Understanding who is at risk of developing PTSD allows for timely clinical intervention and possible prevention.

## Introduction

Posttraumatic Stress Disorder (PTSD) is characterized by symptoms of re-experiencing, avoidance, emotional distress, and heightened physiological arousal following exposure to a traumatic event. 65-89% of all individuals will face a significant traumatic or stressful life event (TSLE) in their lifetime^1–3^, but only 10-30% will develop PTSD^4,5^. Understanding the relative roles of trauma and genetics in PTSD diagnosis will be key to identifying who might develop PTSD following trauma. Therefore, in this study we combine measures of TSLEs with genetic measures such as polygenic risk scores (PRS) and genome-wide association study (GWAS) results using a large hospital-based biobank.

The genetic etiology of PTSD varies according to gender^6–8^, and index trauma type^9–11^; consequently, studies that examine relatively homogeneous cohorts – i.e. individuals exposed to only one specific type of trauma^12–18^ – have identified genome-wide significant associations that have not yet been identified using meta-analytic approaches across trauma types. Reducing cohort heterogeneity by characterizing trauma is imperative to advancing PTSD genetic discovery. Characterizing individuals according to a single index trauma may help, but could fail to fully capture trauma histories among cohorts. Single index trauma characterization cannot account for degree or chronicity, and may fail to identify truly causal traumas within the cohort. Therefore, in this study we combine various types of Electronic Health Record (EHR)-derived TSLEs to construct individual-level trauma scores to capture chronicity and degree of trauma exposure.

The benefits of using EHR data are numerous. First, data is readily available and does not have to be collected, reducing the time it takes to conduct a study^19^. Second, EHR encompasses a more diverse range of individuals, potentially including all patients entering the hospital system^20^. Third, the range of TSLEs available may allow us to identify unexpected relationships with PTSD or other psychiatric disorders. For example, interactions between polygenic risk score (PRS) and childhood trauma (CT) explain more variability in Major Depressive Disorder (MDD) than PRS alone^37,21,22^, and CT has been linked to bipolar disorder^23,24^, schizophrenia^24^, and cognitive decline^25^.

In this study, we used EHR data from the Mount Sinai Bio*Me*™ biobank (Bio*Me*™) to identify 22 TSLEs, and tested for associations with PTSD. We found significant associations between individual TSLEs and groups of TSLEs with PTSD. Additionally, we found that multiple TSLEs interact to increase disease risk. We also discovered significant interactions between TSLEs and PTSD-PRS, suggesting a that trauma interacts with common variants to increase disease risk. To further support this finding, we performed a PTSD-GWAS and looked at the interaction between significant SNPs and TSLEs, which increased the variance explained in PTSD. We hope that these findings will allow better understanding of the complexity of trauma in PTSD and improve patient intake, diagnostics, and future care.

## Methods

### Participants from the Mount Sinai Bio*Me*™ biobank

Our study includes 31,704 individuals from Bio*Me*™, with matched genotype^26^ and EHR data. Bio*Me*™ is highly diverse (self-reported ancestry: HA, 35%; EA, 34%; AA, 25%; Other 6%), with an average age of 59.84 (SE=17.85), and an average EHR records length of 10.4 (SE=4.2) years. A total of 58.7% of the sample self-identify as women; 39.2% as men; 2.1% as non-binary.

### Defining TSLEs

Since our sample has not been collected for a particular study of trauma, but rather is an existing patient biobank, trauma phenotypes may not have been equally assessed across the sample. For example, patients already seeing a psychiatrist are more likely to be asked about trauma.

Therefore, we sought to identify TSLEs that might be more generally captured in the EHR, such as serious illness and self-actualization traumas, and additionally identified positive, negative, and null trauma controls. We selected positive controls, including codes that may be proxies or pre-cursors to PTSD diagnoses (e.g. grief); negative controls that we expect to be protective (e.g., marriage), and null controls that we expect to be unassociated with PTSD or TSLEs.

We included only TSLEs occurring in at least 10 patients. We divided TSLEs into five literature-defined groups^27^ (**Table 1**): personal (e.g. domestic violence), exposure to systematic discrimination (e.g., racism, homophobia, inferred by proxies), self-actualization (e.g. homelessness), physical survival (e.g., serious illness) and interdependence (affecting a direct relative).

### Defining PTSD

We used a broad case-definition of PTSD which included all International Classification of Disease (ICD)-10 codes (≥ 1 count) under the F43 group classification. Controls included all individuals with zero counts under the F43 group classification.

### Trauma contribution to PTSD

We tested for associations between each TSLE and ICD-code defined PTSD using a linear regression model, corrected for age. We applied a Bonferroni correction to establish significance, correcting for the number of TSELs and controls tested.

### Quantifying trauma by individual

In order to assess the ‘amount’ of TSLEs experienced by each individual within our biobank, we created three trauma scores (TRS): first, a simple aggregate trauma score (TRS_SUM_); a score weighted by frequency of each TSLE for each individual (TRS_FREQ_); and a score derived from a joint elastic next model (TRS_EN_). To create our TRS_EN_, we applied a 10-fold cross-validation approach using the “cvglmnet” function in *glmnet* elastic net regression package in R^28^. We define each of our trauma scores as follows (Eq. 1):

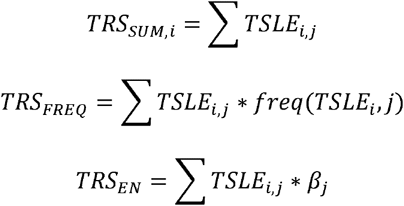

Where:

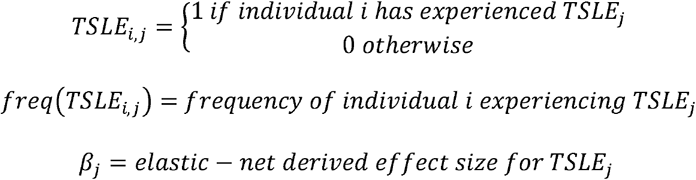

*Equation 1. Trauma risk scores derived from Electronic Health Records data*.

### PRS were calculated for PTSD ancestries

We tested the PTSD PRS of specific ancestries with N >100 PTSD cases; these were AA-PTSD (R^2^=0.02, P_t_=2.5×10^−4^, p=0.011), EA-PTSD (R^2^=0.07, P_t_=0.028, p=0.142) and HA-PTSD (R^2^=0.049, P_t_=0.32, p=0.028). PRS were derived from the largest Psychiatric Genetics Consortium (PGC) PTSD GWAS^6^ separately for each ancestry. We fit the best model for each ancestry using PRSIce-2^29^. Variance explained was calculated correcting for age.

### PRS were tested for interactions with trauma

We tested for interactions between trauma and PTSD PRS using a linear model, correcting for age (Eq. 2).

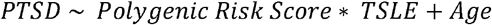

*Equation 2. Linear regression to determine which TSLEs interact with PRS for PTSD*.

### GWAS meta-analysis of all three ancestries

We performed three ancestry-specific PTSD GWAS, in AA, HA, and EA samples within Bio*Me*™; we excluded other ancestries with <100 cases. We defined PTSD using a broad definition in line with PGC-PTSD GWAS^30^, resulting in a total sample size across all three cohorts of 1,990 PTSD patients and 28,478 controls. SNPS that had a MAF of <1%, Hardy-Weinberg equilibrium P>0.001 or call rate <98% were excluded. GWAS were calculated using the first 10 principal components and age as covariates. Meta-analysis across all ancestry groups was performed using an inverse-variance-weighted fixed effects model with METAL^31^.

### Finemapping of GWAS meta-analysis results

We performed finemapping on all GWAS loci including at least one SNP reaching p<1×10^−5^ in our meta-analysis to identify putatively causal SNPs. Finemapping was performed using PolyFun^32^ in combination with FINEMAP^33^. Lead SNPs were analyzed for causal variants contained in the locus 500kb upstream and downstream from start site. Casual variants were selected based on a Posterior Inclusion Probability (PIP) that constitutes a total (PIP=1.0) based on the top two variants in the locus (PIP>0.40 for all variants).

## Results

### Identification of traumatic and stressful life events in the Mount Sinai Bio*Me*™ biobank

We identified 23 traumatic and stressful life events (TSLEs) occurring in >10 individuals within Bio*Me*™ (**Table 1A**). TSLEs were split into 5 categories: personal, exposure to systematic discrimination, self-actualization, physical survival, and interdependence^27^. In addition, we selected nine positive negative and null controls.

22/23 TSLEs were significantly associated with PTSD (p<1.61×10^−3^). The top three associations included surgery (*β*=0.069, p=2.95×10^−120^), HIV+ (*β*=0.145, p=1.37×10^−103^), and female sex (*β*=0.033, p=6.80×10^−31^; **Table 1A**).

Using a linear regression model weighted by an individual’s frequency of exposure to a trauma, we identified one additional TSLE that was significantly associated with PTSD: heart attack (*β*=0.012, p=8.47×10^−4^; **Supplementary Table 1**). Otherwise, TSLE significance and effect size in association with PTSD was similar between the two models.

In line with our initial hypotheses, we identified positive associations between PTSD and all four positive controls, and negative associations between PTSD and marriage. Getting a flu vaccine (null control) was not significantly associated with PTSD, as expected. Most significantly, psychotherapy was associated with an increased risk of PTSD (*β*=0.504, p=2.56×10^−75^), while marriage was most significantly protective against PTSD (*β*=-0.027, p=3.99×10^−18^; **Table 1A**). The other negative/null controls were small in effect size, but significantly positively associated with PTSD (**Table 1A**).

Next, we hypothesized that specific combinations of TSLEs may represent a greater risk for PTSD. Therefore, we tested for interaction effects between all 21 TSLEs, and identified nine significant interactions. The most significant was being female: HIV (*β*=0.013, p=8.9×10^−11^); i.e., women who are infected with (or test positive for) HIV are at greater risk of PTSD than men infected with HIV (**Table 1B**). We found that pairs of TSLEs with significant interactions tended to span categories, rather than coming from the same category (**Figure 1A**).

**Figure 1.**
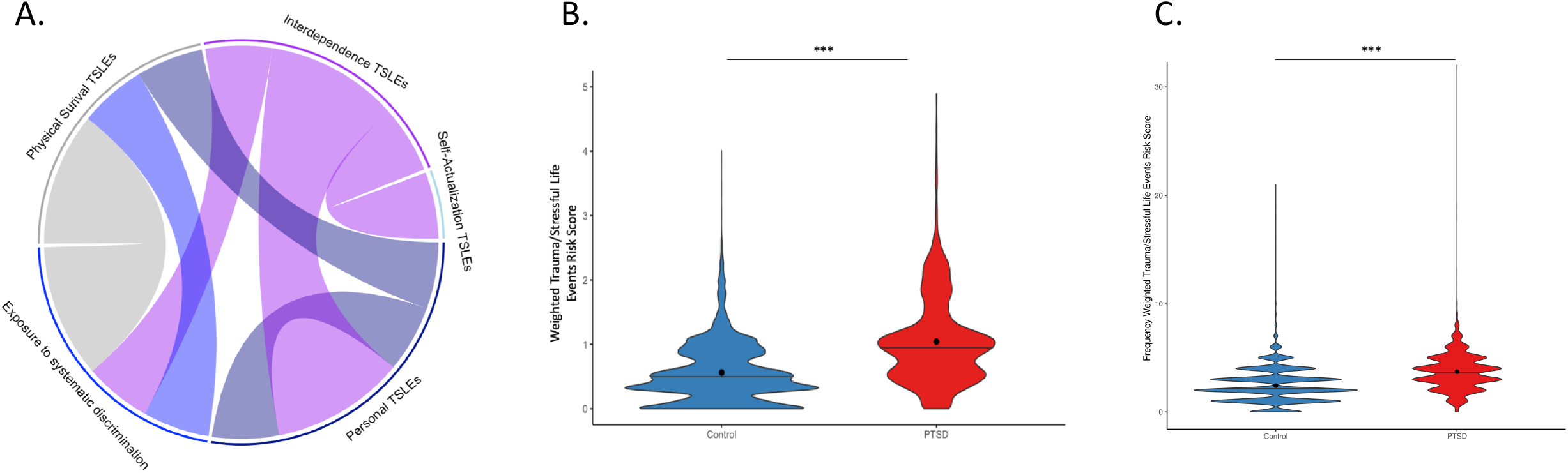
Trauma risk score (TRS) category interactions and averages among PTSD cases and controls. (A) Interactions between Traumatic/Stressful Life Events (TSLEs) of TRS categories. (B) Average TRS (weighted by elastic net regression beta values) between PTSD cases and controls and (C) average TRS (weighted by frequency of trauma exposure) between PTSD cases and controls.

### TSLE risk score measures the association between disease risk and TSLEs

We tested whether aggregate TSLE exposure increases risk for PTSD beyond the risk posed by single TSLEs and, if so, whether the number of TSLEs, frequency of each TSLEs, or specific combinations of TSLEs increase the risk further. To do so, we created three trauma risk scores: a simple aggregate trauma score (TRS_SUM_); a score weighted by frequency of each trauma for each individual (TRS_FREQ_); and a score derived from a joint elastic next model (TRS_EN_).

The total number of TSLEs experienced (TRS_SUM_) was significantly correlated with PTSD diagnosis (p=2.57×10^−229^), explaining ∼3.3% of phenotypic variance within our sample. TRS_SUM_ was significantly higher among PTSD cases compared to controls (TRS_SUM_ average = 2.534 [SD =1.15] vs. 1.718 [SD=1.07]; p=3.353×10^−173^). Similarly, the frequency of each TSLE (TSLE_FREQ_) was significantly associated with PTSD (p=2.56×10^−270^), explaining ∼4% of the phenotypic variance, and was significantly higher in cases compared to controls (TRS_FREQ_ average = 3.707 [SE=1.79] vs. 2.415 [SE=1.56]; **Figure 1C**).

Finally, TRS_EN_ explained 5.44% of phenotypic variance in PTSD (p=1.0×10^−300^), substantially more than either of the other scores. Like the previous TRS, TRS_EN_ was significantly higher among cases compared to controls (average = 1.04 (SE=0.673) vs. 0.562 (SE=0.461); p<1.4×10^−72^). No single category of TSLEs drives this effect; in total, our TRS_EN_ includes 17 TSLEs **(Supplementary Table 2)**, including TSLEs from each of our five categories. Constructing TRS_EN_ using each category individually also produces significantly different TRS_EN_ scores between cases and controls for every group except self-actualization TSLEs (**Supplementary Figure 2**).

### Contribution of TSLEs to the prevalence and severity of other psychiatric disorders

We performed a phenome-wide association study (PheWAS) to identify other psychiatric traits associated with TRS_EN_ (**Supplementary Figure 3**). We identified 37 psychiatric and substance-use traits, and 25 neurological traits significantly associated with TRS_EN_ at Bonferroni significance (**Table 2**): including anxiety disorders (PheCode 300.1, OR=1.570, p=8.91×10^−305^), adjustment reaction (PheCode 304, OR=2.282, p=2.46×10^−206^), and PTSD (PheCode=300.9, OR=1.888, p=4.99×10^−107^). Similarly, we identified 27 psychiatric and substance-use traits, and 25 neurological traits significantly associated with the TRS_FREQ_ at Bonferroni significance (**Supplementary Table 3**). There were 16 disorders that were differentially associated with TRS_FREQ_ and TRS_EN_, including pain (PheCode 338, OR=1.492, p=6.25×10^−180^), lack of coordination (PheCode 350.3, OR=1.27, p=1.43×10^−10^) and meningitis (PheCode=320, OR=1.327, p=1.29×10^−8^) for TRS_FREQ_ and memory loss (PheCode 292.3, OR=1.489, p=5.49×10^−61^), dementia (PheCode 290.1, OR=1.299, p=7.98×10^−17^) and mild cognitive impairment (PheCode 292.2, OR=1.428, p=3.71×10^−13^) for TRS_EN_.

We also performed a PheWAS on each of our TSLE categories to measure their phenotypic associations (**Table 2**). Except for the negative controls, each category was significantly associated with psychiatric disorders. For systematic discrimination-TSLEs and positive controls, the most highly-associated psychiatric disorder was anxiety disorders (PheCode=300, OR=2.335, p=1.87×10^−58^; OR=3.857, p=9.72×10^−294^). Interdependence- and physical survival-TSLEs were most highly associated with mood disorders (PheCode=296, β=0.398, p=1.81×10^−54^; OR=1.898, p=1.42×10^−222^). Personal-TSLEs were most highly associated with PTSD (PheCode=300.9, OR=3.013, p= 3.41×10^−12^), and self-actualization-TSLEs were most highly associated with MDD (PheCode=296.22, OR=3.456, p=1.61×10^−20^; **Supplementary Figure 4**).

### GWAS meta-analysis reveals PTSD SNPs that interact with TSLEs

A key motivator of our study was to investigate whether including trauma in our studies might more deeply elucidate the genetic architecture of PTSD^12^. Therefore, we conducted a GWAS within our sample, following PGC broad PTSD diagnostic definitions^7^, and refined our associations using FINEMAP. As expected given our modest sample size, we did not identify any genome-wide significant associations (**Supplementary Figure 5**).

Based on finemapping results, we selected 18 SNPs (p<1×10^−5^), and tested for interaction effects with individual TSLEs and TRS. We found six Bonferroni significant SNP x TSLE or SNP x TRS_EN_ interactions (p<7.51×10^−5^; **Table 3**), and 10 Bonferroni significant SNP x TSLE_FREQ_ or SNP x TRS_FREQ_ interactions (p<1.33×10^−4^). Of these interactions, five increased in significance compared to the GWAS and reached genome-wide significance: rs189796944 x TRS_EN_ and TRS_FREQ_ (*β*>0.060, p<9.04×10^−9^), rs189796944 x PS-TRS_EN_ and PS-TRS_FREQ_ (*β*>0.127, p<4×10^−8^), and rs113282988 x TRS_FREQ_ (*β*>0.056, p<3.28×10^−9^; **Table 3**).

### PRS elucidate more GxE interactions with PTSD

Finally, we tested whether including TSLEs and TRS in concert with PRS might help to clarify individual-level risk for PTSD. We tested whether PRS significantly predicted PTSD alone and in concert with TSLEs and TRS, in three ancestry-specific groups within our biobank (European, EA; African-American, AA; Hispanic American, HA).

First, we compared the amount of variance explained when including (i) PRS alone (ii) TRS alone and (iii) an interaction between PRS and TRS (**Supplementary Table 5**). For all ancestries, we found a significant increase in variance explained when including the interaction term, compared to PRS alone (p<2.6×10^−102^). However, in all circumstances, TRS alone explained the most variance (R^2^=3.3-5.4%, p<2.56×10^−270^).

Next, we considered more specific associations between PRS and TSLEs and TRS. In our European (EA) cohort, we found significant interactions between PTSD-PRS and personal-TSLEs (TRS_EN_; *β*=978.55, p=4.71×10^−4^), self-actualization-TSLEs (TRS_EN_; *β*=1045.52, p=3.66×10^−3^), and positive controls (**Table 4**). For individual TSLEs in EA, we found interactions between PTSD-PRS and domestic violence (*β*= 591.53, p=1.07×10^−3^), and between PTSD-PRS and sexual assault (*β*=207.74, p=1.80×10^−3^). We found a significant interaction with frequency of personal-TSLEs and EA-PTSD-PRS, both overall (*β*=264.31, p=4.09×10^−3^) and for 4 specific TSLEs: psychotherapy, heart attack, work stress, and sexual assault (*β*>207.43, p<6.58×10^−4^; **Table 4**). We also observed a significant interaction between TRS_EN_ and PTSD-PRS for in our African-American cohort (*β*=14.24, p=2.71×10^−3^).

Finally, we considered whether TRS and PRS applied in concert might provide more diagnostically useful risk-stratification. To do so, we compared the proportion of cases and controls falling in the top decile to (i) the bottom decile; and (ii) the bottom 9 deciles, using (i) PRS alone; (ii) TRS alone; (iii) PRS and TRS in concert. Across all ancestries, individuals in the top decile of TRS risk were significantly more likely to be PTSD cases, compared to the bottom decile (OR>7.87, p<1.37×10^−36^; **S. Table 5**), and the bottom nine deciles (OR>3.75, p<8.41×10^−43^). Individuals in **both** the top TRS **and** PRS deciles were more significantly enriched for cases in AA and HA populations (OR>8.50, p<2.62×10^−8^). Although numbers are small, the same pattern holds when considering the top quintile (i.e., top 20%; **S. Table 5B**)

## Discussion

In addition to DSM-5 recognized PTSD-associated traumas (e.g. sexual assault, domestic violence, grief), we also discovered a large number of atypical TSLEs associated with PTSD, such as systematic discrimination measures (e.g. racism, sexism, homophobia), stress from work, finances, and other physical health stressors such as cancer or experiencing a heart attack. We examined whether these interactions occur across or within TSLE categories to address whether two similar TSLEs contribute more risk for PTSD than two independent TSLEs. We found that while TSLEs do interact, the majority of interactions are between different TSLE categories, rather than within the same category. Interaction of these TSLEs leads to a higher risk of developing PTSD, and thus led us to investigate the risk and implications of experiencing multiple traumatic events.

To expand on the idea that multiple TSLEs lead to a higher risk and more severe PTSD symptoms, we constructed an aggregate trauma risk score (TRS) of TSLEs for an individual and found that using elastic net-derived effect sizes predicts more phenotypic variance in PTSD than an unweighted model, exposure-counts weighted-model, or PTSD-PRS. On average, individuals with PTSD experience a higher number and frequency of TSLEs.

The association between trauma exposure and risk of developing a psychiatric disorder has been demonstrated in psychiatric disorders other than PTSD, such as MDD^22,34–37^, substance abuse^38– 40^, and suicidality^38^. However, the influence of trauma-specific factors (type, chronicity, etc.) on disease risk has not been well studied for the majority of psychiatric disorders. Our PheWAS analysis suggests that trauma is significantly associated with many psychiatric disorders. As such, psychiatric disorder studies may benefit from the incorporation of trauma information into the study design. To accomplish this, trauma screening questionnaires should be implemented as a standard tool in all psychiatric disorder studies.

Two loci reached genome-wide significance in interactions with our TRS: rs113282988 (an intronic variant in *ITGAE*) and rs189796944 (an intronic variant in *DPYSL2). DPYSL2*, also known as *CRMP2*, is involved in facilitating neuron guidance, growth and polarity^41–43^. It is also implicated in multiple neurological disorders^44^ including Alzheimer’s disease^45,46^, schizophrenia^47–52^, and alcohol-dependence^53^. *ITGAE* canonically mediates the adhesion of intra-epithelial T-lymphocytes to epithelial cells and is involved in intra-tumoral immunity^54–56^, but it has also been implicated in Parkinson’s disease^57^ and Attention-deficit/hyperactivity disorder (ADHD)^58–60^.

We found that our TRS significantly improves the variance explained in PTSD over PRS. We see significant interaction between PRS and TRS (p< 1.93×10^−49^), suggesting that common variants, in concert with TRS, increase PTSD risk. In addition to TRS interactions, we found interactions between PRS and domestic violence, and sexual assault in EA. It has been well documented that these TSLEs can individually lead to PTSD^61–65^, but this is the first study to specifically test whether the multiplicity of experiencing these TSLEs can lead to a higher risk of developing PTSD. Additionally, we found that individuals who into the top decile (or quintile) of both TRS and PRS were significantly enriched for PTSD (OR 4.18-10.05). In both African American and Hispanic American cohorts, individuals in the top TRS and PRS decile were more likely to have PTSD than individuals in the top TRS decile alone; however, this was not the case for Europeans. We hypothesize that this may be due to lower overall levels of trauma experienced in this population; it is possible that individuals who do report trauma are more likely to do so because of a PTSD diagnosis, increasing the predictive accuracy of the TRS. In line with this, the OR for PTSD cases in the top vs. bottom TRS decile was substantially larger for European Americans (OR=12.68) than for African (OR=9.62) or Hispanic Americans (OR=7.87).

There are three potential reasons why our TRS significantly outperforms the PTSD-PRS: (i) the model for TRS is potentially overfit, (ii) the relationship between TRS and PRS in controls, and/or (iii) our TRS itself might be more heritable than PTSD. Our TRS was designed using trauma exposures within our own cohort, and while we used elastic net regression with ten-fold cross validation to attempt to minimize statistical bias, the model may potentially be overfit. Additionally, at lower PRS, the risk of developing PTSD is two-fold lower in individuals with a high TRS compared to a low TRS (**Supplementary Figure 6**). This suggests a potential genetic resilience in individuals with a low PRS. In controls exposed to multiple traumas, individuals with a low PRS are more protected from developing PTSD than those with a high PRS. This interaction may also be moderated by gene expression or epigenetics. Last, we cannot rule out the possibility that our TRS itself is heritable. In a recent PTSD genome-wide by environment interaction study (GWEIS) study, authors found that their unweighted, quantitative trauma risk score had large genetic overlap with PTSD and reduced PTSD heritability by 31% when adjusted for trauma^66^. Similarly, combat^30,67^ and other traumas^68^ have been reported to be heritable.

One significant caveat of this study is the lack of adequate sample size for our GWAS and PRS. We did not reach genome-wide significance in the meta-analysis, and our PRS were very low compared to efforts from the PGC, which found SNP-heritability to be 5-20%, varying by sex^69^.

The diversity and breadth of Bio*Me*^*™*^ gave us a unique opportunity to understand the role of trauma in a clinical population. However, EHR-derived data has some limitations: records are easily obtainable and searchable, but they are not curated for the purposes of this study. We are limited in the number of TSLEs that we can test due to recorded availability and low volume of cases. We chose to exclude any TSLE that had <10 cases, due to the potential to highly skew interactions when limited to a small number of individuals. One example of a TSLE that was excluded was childhood trauma (CT), since so few codes are reported (<10), likely indicating under-reporting rather than an absence of the exposure. To include CT would therefore (1) not capture all the individuals with unreported CT and (2) skew the results of interactions with CT to extreme cases only.

Accounting for trauma increases the amount of variance explained in PTSD, compared to considering genetics alone. Specifically, considering aggregate scores of trauma—weighted according to machine-learning approaches—accounts for a >5-fold increase in the amount of variance explained in PTSD. This suggests that trauma is a critical component in understanding one’s risk of developing PTSD, and a thorough trauma history is necessary to help diagnose and treat patients with PTSD. By examining the interactions between trauma and genetics, we can better understand the etiology of PTSD and design better treatments and prevention methods.

## Supporting information

Supplementary Results

## Data Availability

All data produced in the present work are contained in the manuscript.

