## Supplementary Figure Legends for "Disentangling the roles of trauma and genetics in psychiatric disorders using an Electronic Health Records-based approach"

**Supplementary Figure 1. Trauma risk score (TRS) averages for all races (African American, European American, Hispanic American) between PTSD cases and controls.**

**Supplementary Figure 2. Trauma risk score (TRS) category averages between PTSD cases and controls.** TRS weighted by frequency of trauma exposure.

**Supplementary Figure 3. Phenome-wide association study (PheWAS) of a trauma risk score (TRS) and TRS categories weighted by elastic net regression beta values.**

**Supplementary Figure 4. Forest plots of phenotypes significantly associated with the trauma risk score (TRS) phenome-wide association study (PheWAS) and PheWAS of TRS categories.**

**Supplementary Figure 5. Manhattan plot of a posttraumatic stress disorder (PTSD) genome-wide association study (GWAS) meta-analysis in the Icahn School of Mount Sinai Bio*Me^TM^* biobank.**

**Supplementary Figure 6. Deciles of posttraumatic stress disorder (PTSD) polygenic risk scores (PRS), trauma risk scores (TRS), TRS and PRS converged, and the highest decile of PRS or TRS.**

**Supplementary Figure 7. Average time (days) since posttraumatic stress disorder (PTSD) diagnosis to exposure to a traumatic life event or stressor (TSLE).**

**Supplementary Figure 8. Trauma risk score (TRS) averages between individuals with/without a psychiatric disorder diagnosis or visit/no-visit to an Obstetrician-Gynecologist (OBGYN).** (A) TRS weighted by elastic net regression beta values for individuals with/without at least 1 psychiatric disorder. (B) TRS weighted by elastic net regression beta values for individuals without/with at least 1 visit to an OBGYN. (C) TRS weighted by frequency of trauma exposure for individuals with/without at least 1 psychiatric disorder. (D) TRS weighted by frequency of trauma exposure for individuals without/with at least 1 visit to an OBGYN.
