## Supplementary Results for "Disentangling the roles of trauma and genetics in psychiatric disorders using an Electronic Health Records-based approach"

**Supplemental Results**

**Assessment of TSLEs and control phenotypes in relation to TRS and time since PTSD diagnosis**

To gain a more in-depth understanding of each TSLE/control and their relationship to PTSD, we calculated the average time since PTSD diagnosis that each individual was exposed to TSLEs/controls. In total, there were 16 TSLEs and 7 controls that had sufficient cases (>5) and matching time data to report (**Supplementary Figure 7**). For two of our positive controls: grief (average_days_=-64.52, SE=332.05) and stress (average_days_=-70.82, SE=571.67), the average days since PTSD diagnosis was closest to zero days out of all phenotypes (except homelessness: (average_days_=-2.33, SE=341.19), meaning they were likely reported together at the time of the hospital visit. Sleep disorders (average_days_=238.20, SE=1694.64) were farther from zero than the other positive controls, and had a large average standard error. Our negative controls -vaginal birth (average_days_=-1252.00, SE=1246.53), pregnancy (average_days_=402.62, SE=1741.56)- and null controls -physical exam (average_days_=-33.29, SE=2441.82), flu vaccination (average_days_=76.50, SE=717.93)- were less conclusive in their time-dependent relationship to PTSD diagnosis due to their large average standard errors (**Supplementary Figure 7**).

To further understand how TSLEs are recorded and distributed in our biobank population, we used our TRS to assess individuals with psychiatric diagnoses and Obstetrician-Gynecologists (OB-GYN) visits. We compared the TRS between individuals with at least 1 psychiatric diagnosis (using ICD10 codes) and patients without any psychiatric diagnosis (**Supplementary Figure 8**). We found that in both TRS (TRS_EN_, TRS_FREQ_), individuals with at least 1 psychiatric diagnosis had significantly (p<2.2x10^-16^) higher average TRS (TRS_EN_=0.480-1.923, TRS_FREQ_=1.213-4.619) than individuals without a psychiatric diagnosis (TRS_EN_=0.275-1.409, TRS_FREQ_=0.714-3.576). Similarly, we compared individuals with at least 1 visit to an OB-GYN to individuals without an OB-GYN visit (**Supplementary Figure 8**), and found that individuals who visited an OB-GYN experienced a significantly higher average TRS (TRS_EN_=0.838-2.024, TRS_FREQ_=2.057-5.067) than those who never visited an OB-GYN (TRS_EN_=0.259-1.543, TRS_FREQ_=0.713-3.755).
