## Supplementary figures and images for "Disentangling the roles of trauma and genetics in psychiatric disorders using an Electronic Health Records-based approach"

### Supplementary Figure 1

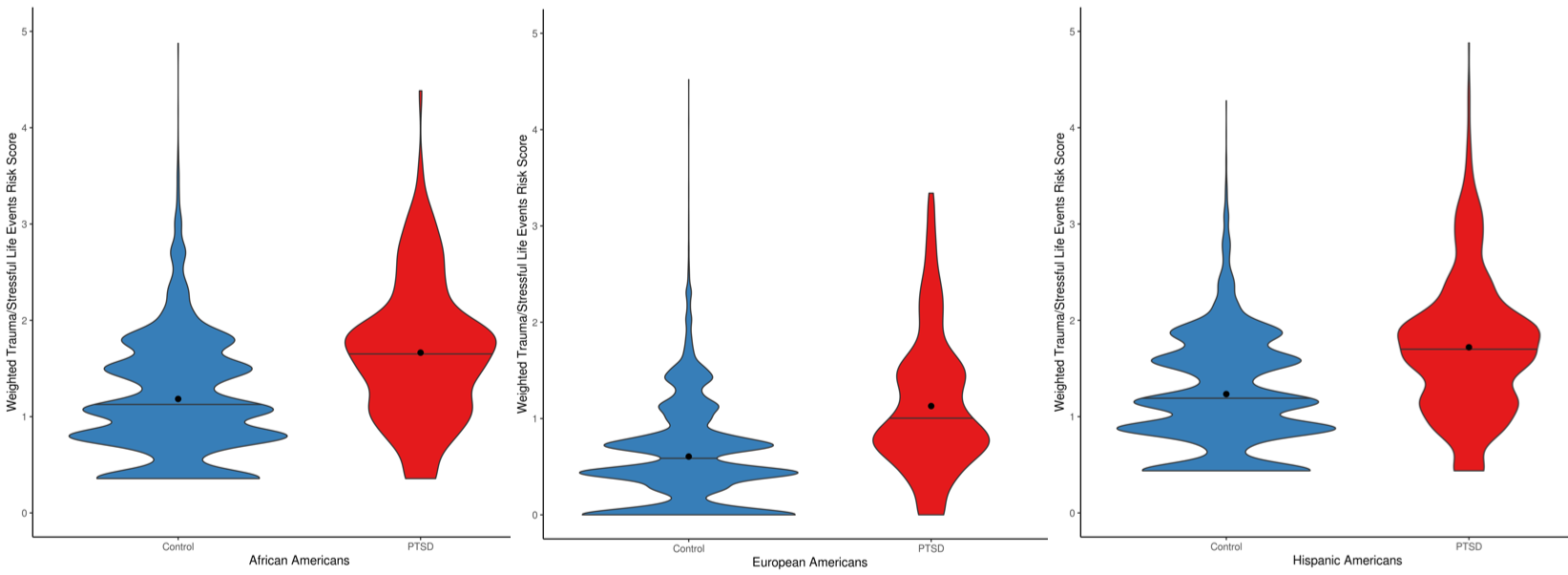

### Supplementary Figure 2

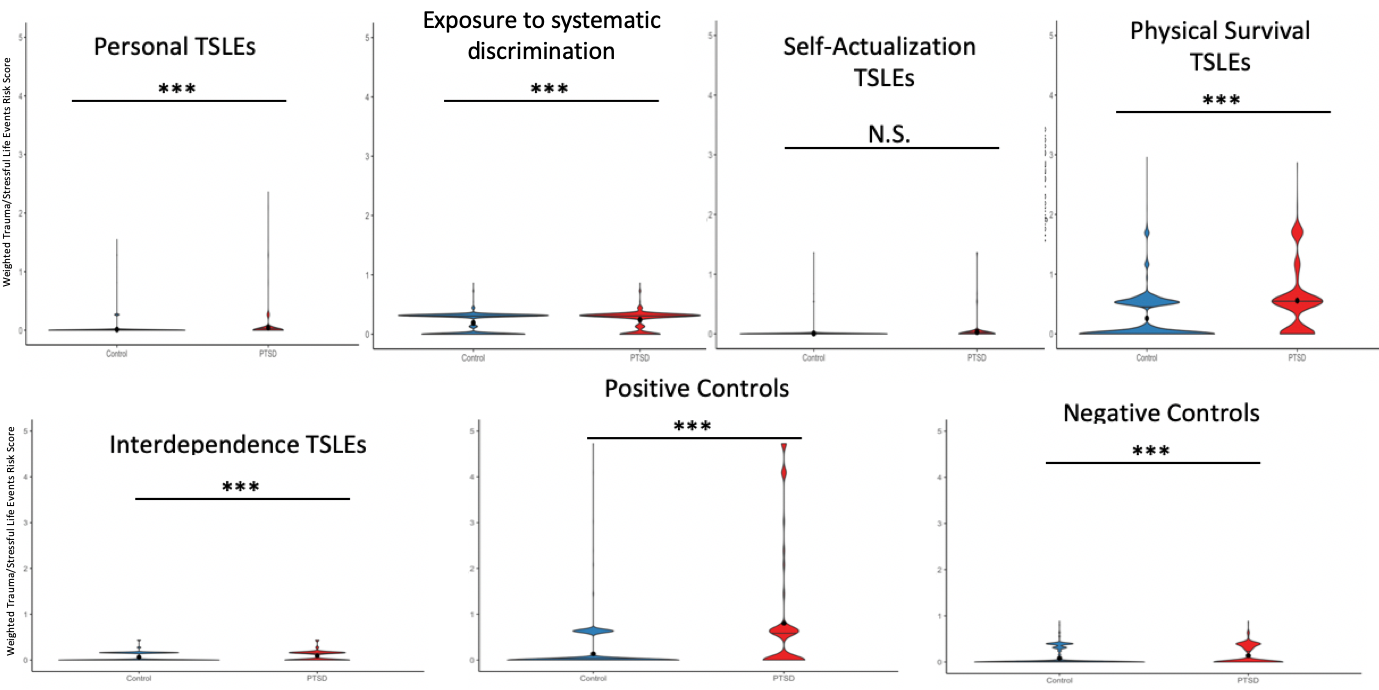

### Supplementary Figure 3

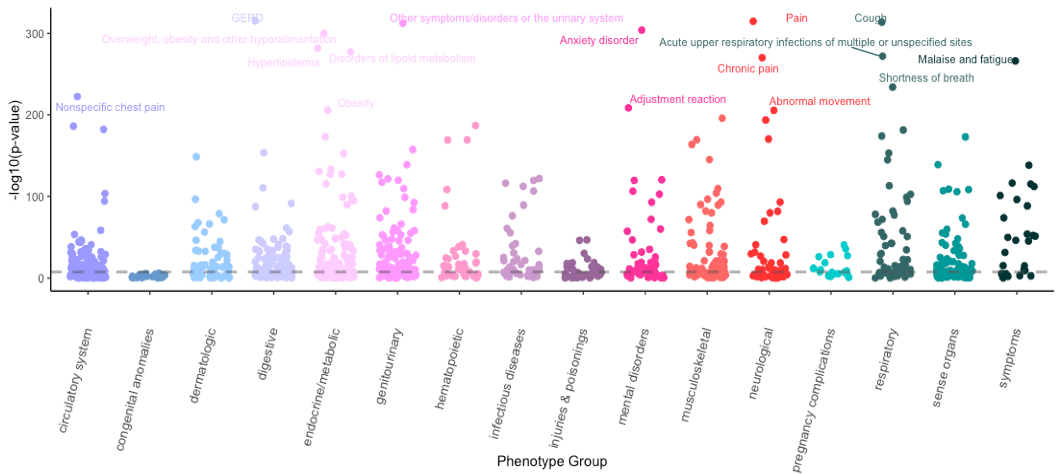

### Supplementary Table 4

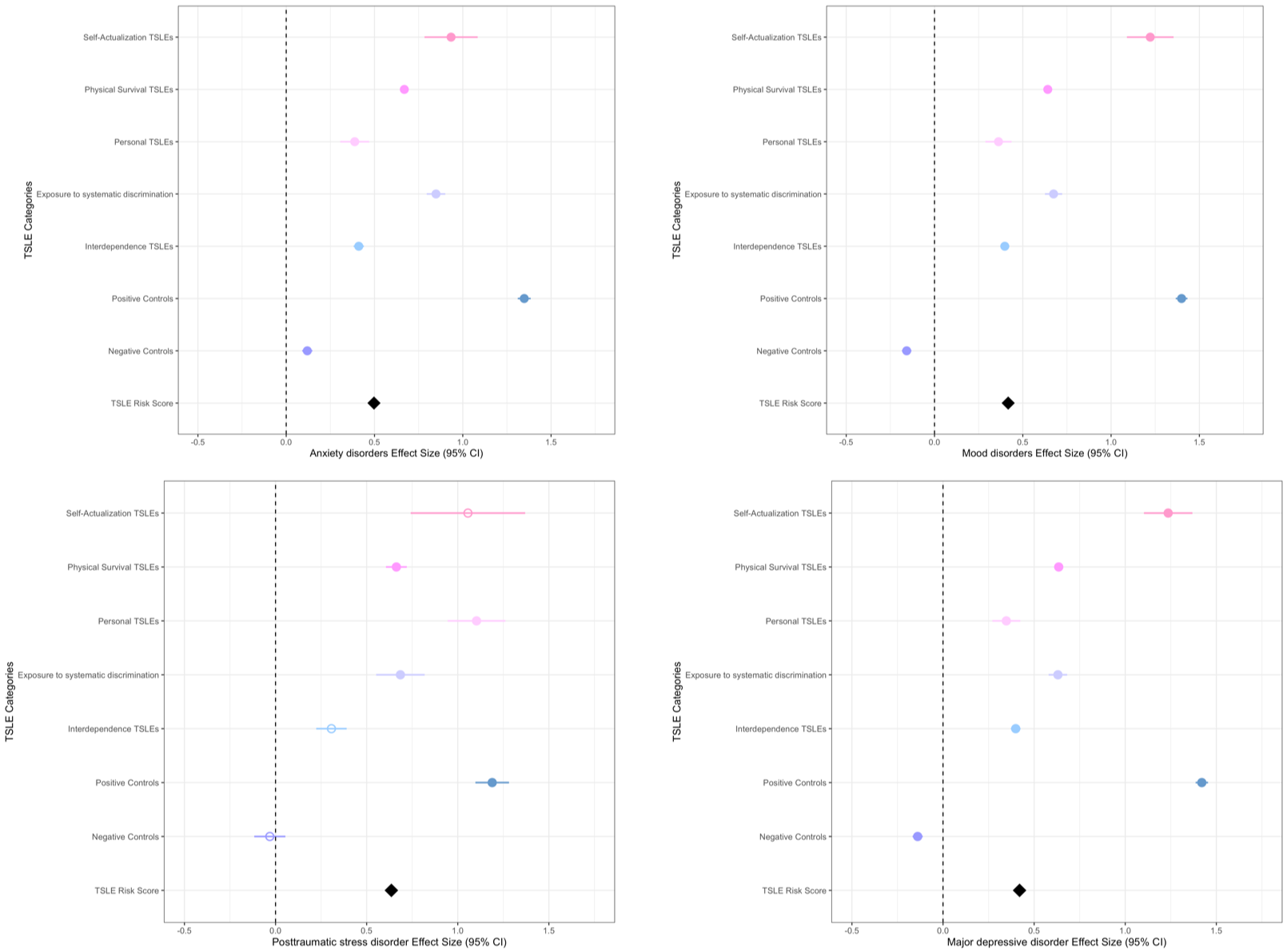

### Supplementary Table 5

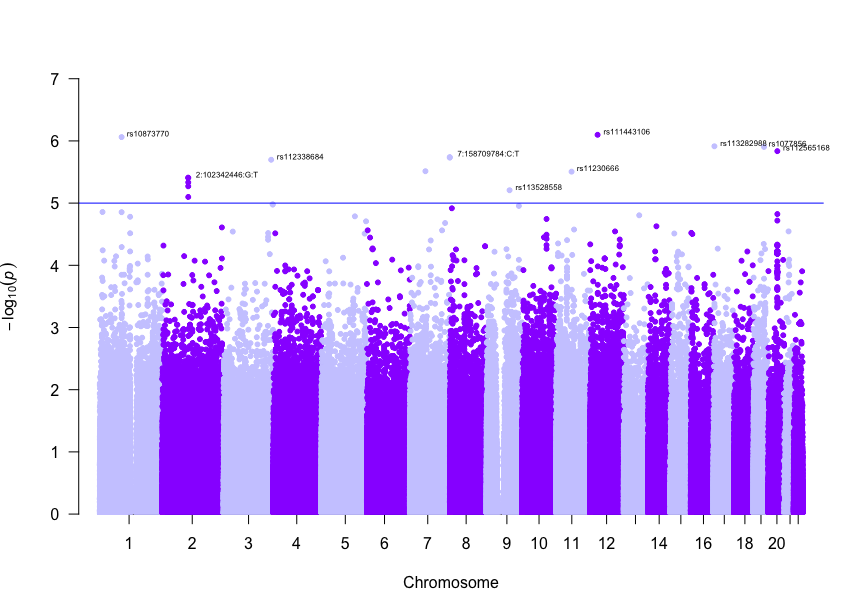

### Supplementary Table 6

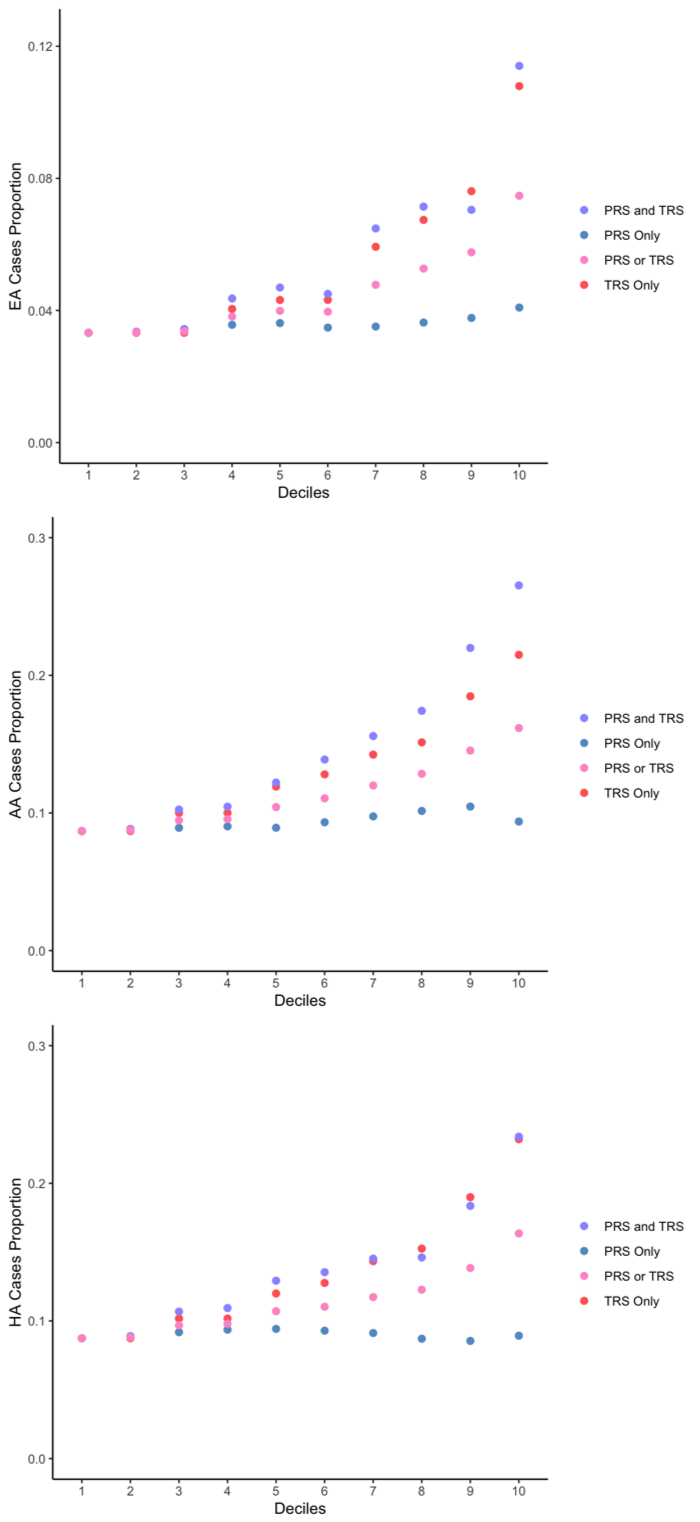

### Supplementary Table 7

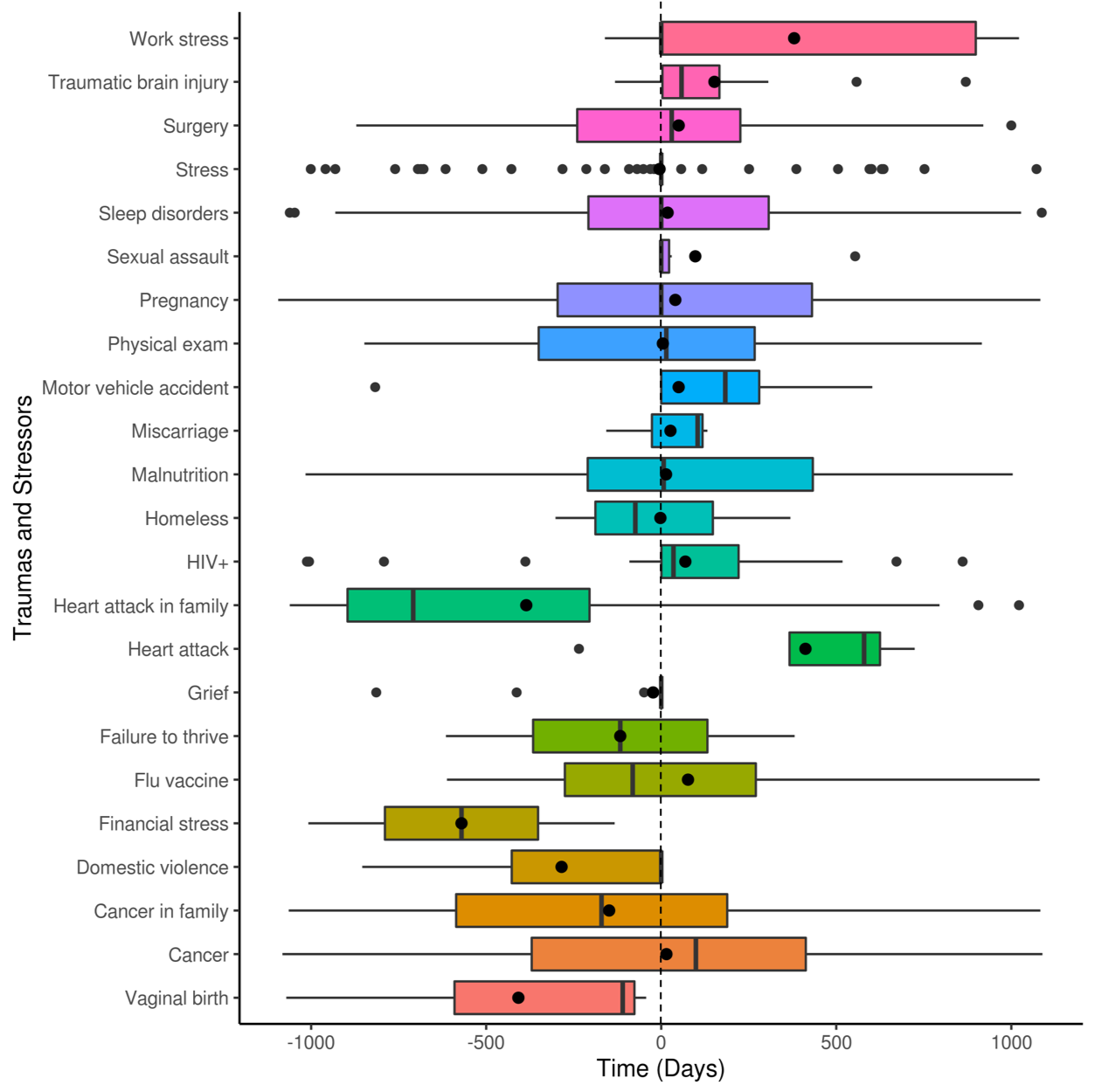

### Supplementary Table 8

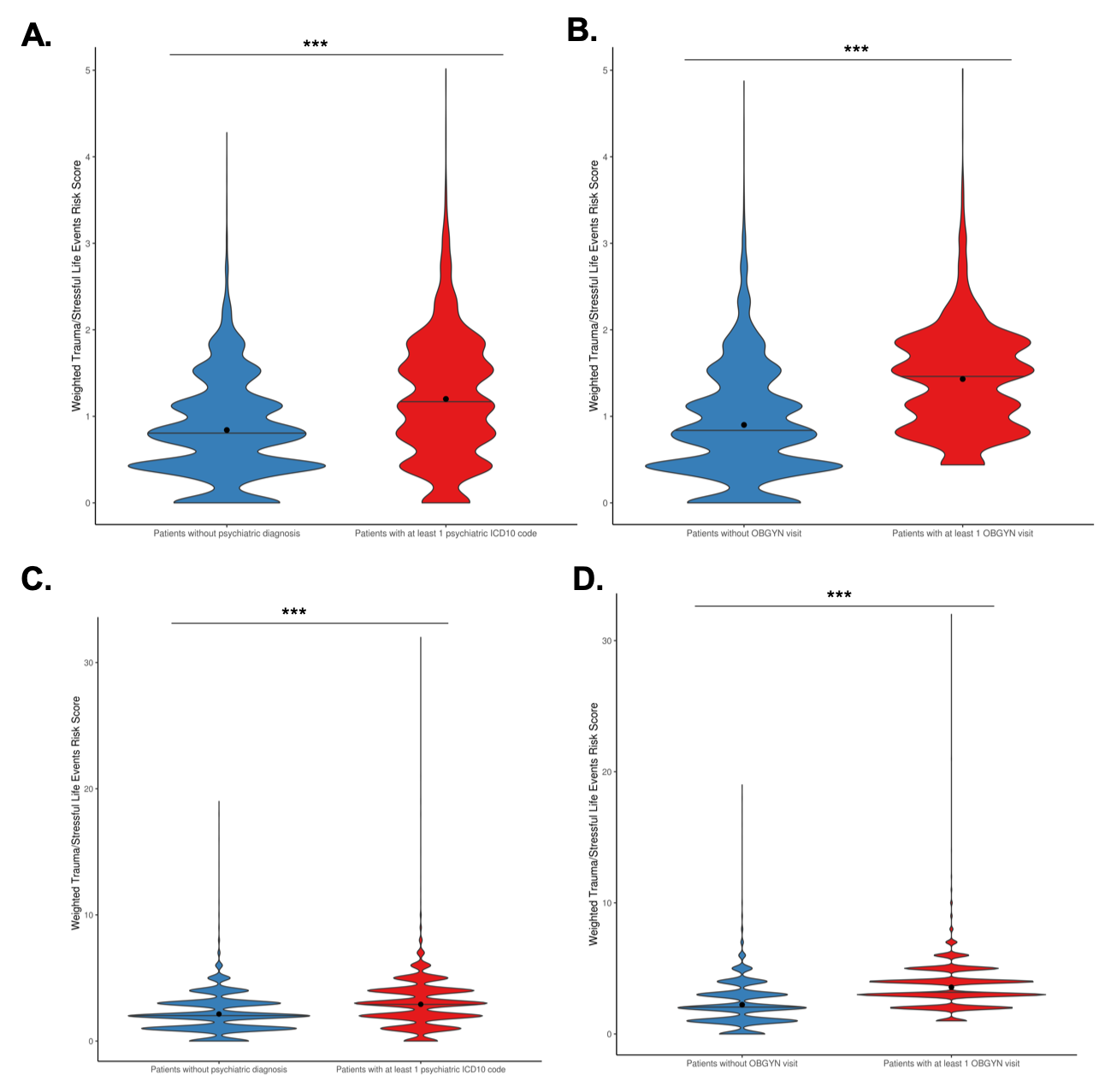
